# Evaluating the adequacy of Prima Covid-19 IgG/IgM Rapid Test for the assessment of exposure to SARS-CoV-2 virus

**DOI:** 10.1101/2020.05.30.20117424

**Authors:** Giulia Di Lorenzo, Paolo Toniolo, Caterina Lurani, Luca Foresti, Chiara Carrisi

## Abstract

The outbreak of the SARS-CoV-2 in early 2020 found health authorities worldwide unprepared to control the pandemic. The adoption of accurate, rapid and inexpensive methods to identify infected subjects in the general population is of paramount relevance for the control of the disease. We evaluated one of the available serological tests, the *Prima Lab* Covid-19 IgG/IgM Rapid Tests, on 739 volunteers. We first assessed the test’s reproducibility by administering it twice on the same day on 104 subjects obtaining and overall score of 93 percent. Since the intensity of the color in the test line regions varies depending on the concentration of Covid-19 antibodies in each sample and that the determination of the positivity depends strictly on the subjective assessment by the reader, after excluding the subjects whose color intensity was too tenuous to be deemed unquestionably positive by the reader the reproducibility increased to 96%. The test would not perform properly for 6 subjects for a very limited overall technical failure of 0.83%. For 138 subjects information was available regarding a previous Real Time PCR nasopharyngeal swab test performed elsewhere. The correspondence of positive results between the two tests was 90.58% (125/138). In spite of some limitation owing especially to the choice of a self selected population sample, we conclude that *Prima Lab* Covid-19 IgG/IgM Rapid Test represents a low-cost, easily applicable and reproducible tool in detecting SARS-Cov-2 diffusion in the general population.

## 2. Introduction

The World Health Organization (WHO) used for the first time the term novel coronavirus in reference to a coronavirus that affected the lower respiratory tract of patients with pneumonia in Wuhan, China on December 29, 2019. WHO has stated that the official name for the novel coronavirus is Covid-19, coronavirus disease 2019, while the reference name for the virus is severe acute respiratory syndrome coronavirus 2 (SARS-CoV-2) [1].

WHO considers essential for the fight against the spread of SARS-CoV-2 to increase the scientific knowledge about the disease and to develop and test new diagnostic methods for the virus, in order to carry out screenings in the population. In fact, all countries that have made extensive use of diagnostic tests for the entire population, regardless of the presence of symptoms, have achieved a drastic drop in infection and in the spread of infection. To date, the only tool officially recognized by the Italian Ministry of Health for the diagnosis of SARS-CoV-2 virus infection is real time RT-PCR method on samples obtained by bronchoscopy, blood or feces. This diagnostic method has limitations related to the cost of reagents and the time required for the complete analysis along with an unacceptably large number of false negatives [2]. Therefore, there is an urgent need to develop accurate, rapid and inexpensive methods to be combined with epidemiological, radiological [4, 5] and clinical data [6] in order to enhance RT-PCR potential to identify infected as well as asymptomatic carriers to prevent virus transmission and assure timely treatment [3].

## 3. Materials and Methods

A total of 739 volunteers participated in the study: 496 (67.1%) healthcare workers, 123 (16.7%) administrative workers and 120 (16.2%) patients. All subjects were asymptomatic when tested. By gender, there were 297 males (40.2%) and 442 females (59.8%). The mean age was 42.9 years old, while the median age was 38 years old.

The study was conducted performing *Prima Lab SA* Covid-19 IgG/IgM Rapid Test, a qualitative membrane-based immunoassay for the detection of IgG and IgM antibodies to Covid-19 in whole blood, serum or plasma samples. All tests have been carried out by a nurse taking approximately 20μL of capillary blood with a sterile lancet, pipetting it into the cassette port and adding two drops of diluent to drive capillary action along the strip. The entire rapid test took about 10 minutes. A total of 844 Covid-19 IgG/IgM Rapid Tests were performed.

104 subjects repeated the Covid-19 IgG/IgM Rapid Test twice on the same day. The second test has been performed by the same nurse puncturing a different finger with a new sterile lancet.

Given the limitations imposed by the local authorities on the availability of RT-PCR swabs, we had to rely on self reported information provided by each volunteers. A total of 149 of 739 subjects reported that they had previously undergone a RT-PCR nasopharyngeal swabs.

## 4. Results

A total of 739 volunteers have been tested using Covid-19 IgG/IgM Rapid Test. Results were divided into: 604/739 (81.73%) C, control negatives, 106/739 (14.34%) IgG positives, 19/739 (2.57%) IgM and IgG positives, 7/739 IgM positives and 3/739 (0.41%) Not valid results.

### Technical Reproducibility

104 Covid-19 IgG/IgM Rapid Tests have been repeated twice on the same day to assess the test Technical Reproducibility. 30 (28.8%) were males and 74 (71.2%) were females.

Results obtained performing two Covid-19 IgG/IgM on the same day have been collected in Table 1.

**TABLE 1:** Covid-19 IgG/IgM Rapid Test Technical Reproducibility

| First Test Results | Test Repetition | Confirmed Test | Not Confirmed Test | Technical Reproducibility |
| --- | --- | --- | --- | --- |
| C, control | 43 | 42 | 1 | 98% |
| C, control, IgG | 52 | 49 | 3 | 94% |
| C, control, IgM | 6 | 4 | 2 | 67% |
| C, control, IgM, IgG | 3 | 2 | 1 | 67% |
| **First Test Results -Total** | **104** | **97** | **7** | **93% +/- 2.7%** |

The Technical Reproducibility for the Covid-19 IgG/IgM Rapid Test is 93%.

Since the intensity of the color in the test line regions varies depending on the concentration of Covid-19 antibodies in each sample and that the determination of the positivity depends strictly on the subjective assessment by the reader, a substantial number of tests were classified as uncertain, or unreliable.

*Prima Lab SA* assumes any shade of color in the test line region should be considered as positive. In order to evaluate how the Technical Reproducibility varies with the color intensity the test results have been divided in reliable and unreliable. Considering only the reliable cluster results the Technical Reproducibility increased to 96% (Table 2).

**TABLE 2:** Covid-19 IgG/IgM Rapid Test Technical Reliable Reproducibility

| First Test Results | Test Repetition | Confirmed Test | Not Confirmed Test | Technical Reproducibility |
| --- | --- | --- | --- | --- |
| C, control | 43 | 42 | 1 | 98% |
| C, control, IgG | 34 | 33 | 1 | 97% |
| C, control, IgM | 3 | 2 | 1 | 67% |
| C, control, IgM, IgG | - | - | - | - |
| **First Test Results -Total** | **80** | **77** | **3** | **96% +/- 2.3%** |

The results of the Covid-19 IgG/IgM Rapid Test evaluation study show a Technical Reproducibility range of 93–96%.

### Technical Failure

Among 844 tests performed in 4 cases the control line did not appear and 3 cases resulted with a strange cassette coloration, maybe due to a bad blood race. The Technical Failure of the test measured is 0.83% +/− 1.4%. Interesting to notice 2/4 of not valid results belong to the same subject, who repeated the test the day after, obtaining another not valid test.

### Covid-19 Rapid Test vs. Nasopharyngeal Swabs

149 RT-PCR swabs results performed by different Italian Reference Laboratories in the period of analysis have been collected. In this study only swabs performed between 28 days after and 28 days prior the Covid-19 IgG/IgM Rapid Test performance have been considered (138/149).

Rapid test results have been compared to RT-PCR assuming when negative (C, control) RT-PCR should be found negative, when IgM positive RT-PCR should be positive and when IgG positive RT-PCR should be both positive or negative. 125/138 (90.58%) Rapid Test have been confirmed by RT-PCR (Table 3).

**TABLE 3:**
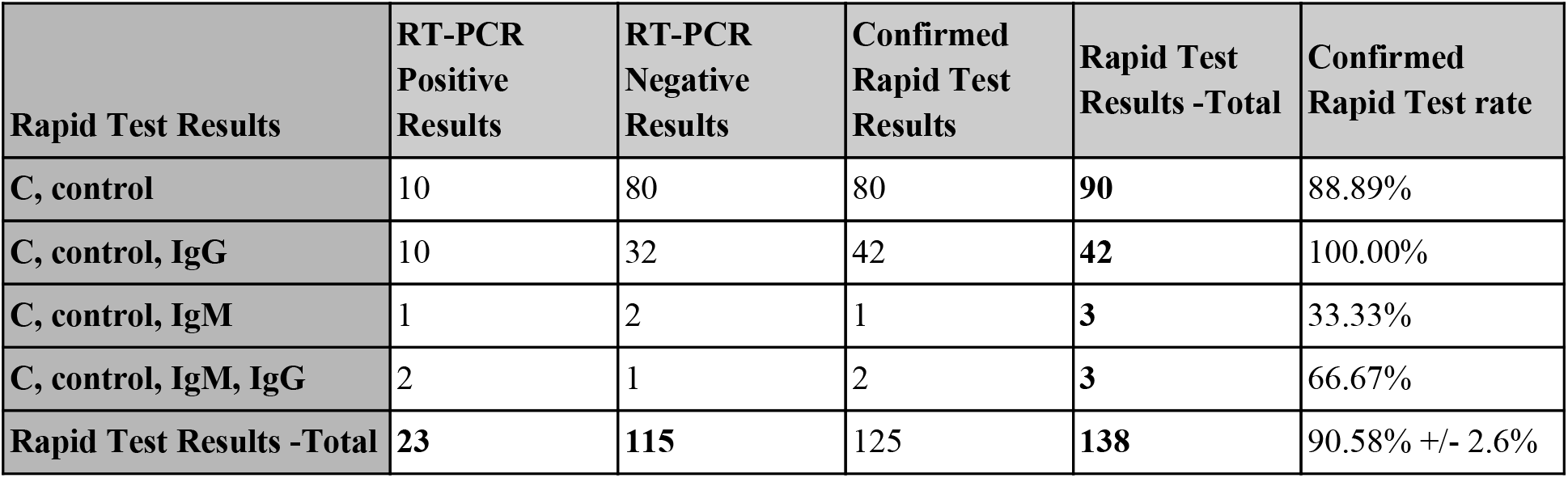
Rapid Test performance vs. Nasopharyngeal Swab

| Rapid Test Results | RT-PCR Positive Results | RT-PCR Negative Results | Confirmed Rapid Test Results | Rapid Test Results -Total | Confirmed Rapid Test rate |
| --- | --- | --- | --- | --- | --- |
| C, control | 10 | 80 | 80 | 90 | 88.89% |
| C, control, IgG | 10 | 32 | 42 | 42 | 100.00% |
| C, control, IgM | 1 | 2 | 1 | 3 | 33.33% |
| C, control, IgM, IgG | 2 | 1 | 2 | 3 | 66.67% |
| Rapid Test Results -Total | 23 | 115 | 125 | 138 | 90.58% +/- 2.6% |

## 5. Discussion

### Technical Reproducibility

The data obtained show a Covid-19 IgG/IgM Rapid Test Technical Reproducibility of 93–96%. This result suggests accuracy is lower than assumed. *Prima Lab SA* claims an accuracy for the Covid-19 IgG/IgM Rapid Test of 92.9% for IgM and 98.6% for IgG, a specificity of 96% for IgM and 98% for IgG and a sensitivity of 85% for IgM and 100% for IgG respectively. According to our data, sensitivity and specificity estimated by the manufacturer should be reduced by a 7–4% factor, corresponding to the reproducibility error highlighted by the analysis.

Technical Reproducibility discrepancy between reliable and unreliable Covid-19 IgG/IgM Rapid Tests results, suggests only reliable results should be considered true. Moreover, every test should be performed at least twice and only the double check results should be considered as correct, in order to get an accurate result.

Recent studies – provided by the producer’s instructions – have shown qualitative membrane-based immunoassay tests are affected by a sensibility and specificity variation for both IgG and IgM antibodies caused by the time between symptoms onset and test performance. The Technical Reproducibility results obtained in this study should be further investigated in correlation with symptoms onset, in order to evaluate if the Technical Reproducibility percentage gets better 16–20 days after the symptoms confirming that this can be considered the best moment for using the Covid-19 IgG/IgM Rapid Test.

### Technical Failure

Invalid results were observed for 7/844, or 0.83% of measurements. *Prima Lab SA* states insufficient sample volume, incorrect procedural techniques or Hematocrit level not falling between 25% and 65% are the most likely reasons for a missing control line.

### Covid-19 Rapid Test vs. Nasopharyngeal Swabs

Covid-19 IgG/IgM rapid test results were compared to RT-PCR for 138 subjects with an observed agreement of 90.58% between the two techniques.

In literature it is known SARS-CoV-2 should be detectable by RT-PCR from 14 days prior to 14 days after symptom onset, with a peak at seven days from symptoms appearance [7]. Anti-Covid-19 antibodies production is expected to start 3–6 days after symptom onset for IgM and 14–21 days for IgG, respectively [8]. This means IgM immunoglobulins should be detectable in the first stage of the disease, when the subject is positive to PCR swab, while IgG immunoglobulins should be detectable in the last stage of disease when the subject become negative to PCR swab.

Since the Rapid Test is most likely to produce negative results in the early stages of infection, it is possible that the 10/90 (11.1%) negative subjects with a positive RT-PCR that SARS-CoV-2 was detectable but antibodies seroconversion has not started yet. Two out of three (66,6%) negative values on IgM and one out of three negatives on IgM/IgG (33.3%) with a negative RT-PCR could be explained as false IgM positive give the low IgM sensitivity of about 85%.

Overall, we found that the Rapid Test appears to be a valid method for detecting both negative SARS-CoV-2 and positive IgG patients. On the contrary, the Rapid Test does not appear to be as effective for the detection of positive or negative IgM subjects and, therefore it should not be performed unless it is followed by RT-PCR.

## 6. Conclusions

At the beginning of the Covid-19 pandemic virtually no information and protocols were available for the clinic management of suspected SARS-CoV-2 patients, but very quickly a number of serological tests became available on the market with the promise to identify people who had or had had the disease. Given the difficulty in securing an adequate supply of RT-PCR kits, we at the Centro Medico Santagostino decided to test a qualitative serological test, the *Prima Lab SA* Covid-19 IgG/IgM Rapid test, to assess the efficacy and the weaknesses of rapid tests.

Over a period of ten weeks, we performed 844 Covid-19 IgG/IgM Rapid tests on 739 volunteers, a convenient sample with no pretense to be representative of the underlying Italian population. The data collected suggests the test has a technical reproducibility of 93–96%, a technical failure of 0,83% +/− 1.4% and an agreement with RT-PCR of 90,58% +/− 2.6%.

The Covid-19 IgG/IgM Rapid test represents a low-cost and easily applicable tool in detecting SARS-Cov-2 diffusion in the general population with the capability to identify quite reliably subjects who have been or have not been exposed to the virus in the recent or distant past, as suggested by the presence or absence of specific IgG. Our data suggest that the test is unlikely to provide adequate information regarding the most recent or current exposure to the virus given the evident unreliability of IgM antibodies detection, which must be conformed by a RT-PCR test.

## Data Availability

The data that support the findings of this study are available from the corresponding author, [C.C.], upon reasonable request.

